# The effect of long-term awareness on active and passive tobacco smokers

**DOI:** 10.1101/2021.05.06.21256782

**Authors:** Serkan Köksoy, Fatih Kara

**Affiliations:** Mehmet Akif Ersoy University, Health Science Faculty, Burdur, TURKEY; Selçuk University, Faculty of Medicine, Konya, TURKEY

**Keywords:** Addiction, Awareness, Carbon monoxide, Respiratory Function Test, Tobacco

## Abstract

**Background and objective:** Tobacco addiction is a major public health problem. Numerous scientific studies have been conducted on the harms of tobacco products. However, the number of intervention studies investigating the effect of long-term awareness of the harm of tobacco products on the Fagerström nicotine addiction test (FTND), carbon monoxide (CO), carboxyhemoglobin (COHb) and respiratory function test (RFT) are limited. Our goal is to investigate the impact of long-term scientific awareness on these parameters.

**Participant and Method:** The study was designed an intervention study on active and passive smoking participants and their control groups. Intervention groups were given seminar program (up-to-date literature information and images on the harms of tobacco products) for eight weeks. Control groups were not given any training on the harms of tobacco products. Intervention groups were measured 8 times (FNBT, CO, COHb and SFT). Control groups were measured only in the first and last week.

**Result:** When comparing the first and last weeks in active smoker intervention group (ASIG), the difference between FTND, FEV1, CO, and COHb parameters was observed to be statistically significant. The highest decrease in ASIG was in CO (60%) parameter and the highest increase was in the FEV_1_(%10) parameter. There was a significant difference between FEV_1_, FVC, FEV_1_/FVC, and CO parameters of passive smokers intervention group (PSIG). The highest decrease in PSIG was in the CO parameter (%65.8) and the highest increase was in the FVC (%10) parameter.

**Conclusion:** Awareness programs may reduce the severity of addiction in active smokers and may help protect passive smokers. In the absence of any awareness, it may not positively change. As awareness increases, positive changes in some vital parameters may be possible. Up-to-date programs and policies are needed to make easy and sustainable awareness of both active and passive smoking.

## INTRODUCTION

The use of tobacco products is a global public health problem that can cause serious health problems and deaths not only in active smokers but also in passive smokers (Mathers and Loncar 2006; Cao et al 2015). In terms of public health, tobacco addiction and tobacco-related diseases are considered a preventable disease (Wang et al. 2018). This preventable health problem causes health problems in every period of life, and especially it affects the individuals in the development age (Bird and Staines-Orozco 2016). Despite scientific warnings and tobacco control programs, tobacco use continues. It occurs in passive exposure in societies where addiction prevalence is high. If this situation cannot be prevented, the chronic disease burden and prevalence related to tobacco smoke may increase (Gentzke et al. 2019). Various epidemiological researches (smokeless air, supply-demand, etc.) have been carried out to find a general solution for both active and passive smoking (Hyland et al. 2012, Vellios et al. 2018). The most feasible and sustainable of these solutions is to raise awareness of the harmful effects of tobacco products and to protect individuals from exposure in every period of life (Nyman et al. 2017, Park et al. 2018). Although a large number of studies have been conducted on the harm of tobacco products, intervention researches that both create long-term awareness and measure of Fagerström Test for Nicotine Dependency (FTND), Carbon monoxide (CO), Carboxyhemoglobin (COHb), and Respiratory Function Tests (RFT) are extremely limited. In order to fill this gap in the literature, up-to-date literature information and visuals about the harm of tobacco products were presented to active and passive smoker intervention groups for eight weeks. It is aimed to compare the findings obtained from the study in terms of both groups.

## PARTICIPANT and METHOD

### General Inclusion Criteria for All Participants

Those who did not have any known health problems, participated in all measurements and seminars, volunteered to take part, non-obese, and between the ages of 18-24 were included in the study.

### Inclusion Criteria for Active Smokers

Having smoked regularly for the past year, having a score of 5 or more on the FTND, smoking at least 20 cigarettes a day, meeting the general inclusion criteria.

### Inclusion Criteria for Passive Smokers

Having lived continuously in an environment (home, dormitory, etc.) with an active smoker (someone smoking at least 20 cigarettes per day and with a score of 5 or more on the FTND) for one year previously, meeting the general inclusion criteria. The participants were informed about the presentations, measurements (RFT, CO) and the duration of the study. A voluntary informed consent form was read to those who agreed to participate in the study.

### Determination of the Sample Size

The sample size of the research was calculated according to subgroups with the G*Power program (Faul et al 2007). The study was designed as two main groups and their two subgroups. The number of samples was calculated according to the subgroups (Effect size: 0.5, α: 0.05, power: 0.80). It was calculated that at least 28 participants must be present in each subgroup to reach 0.80 power.

### Design of the Study

The study was designed an intervention study. The intervention and control groups were separated using the true random number generator (www.random.org). An eight-week presentation program was given to the intervention groups but was not given to the control groups. Intervention groups were measured for each week. The control groups were measured only in the first and eighth weeks. The data obtained from the intervention groups were compared with within group and between group comparisons (Figure 1).

**Figure 1:**
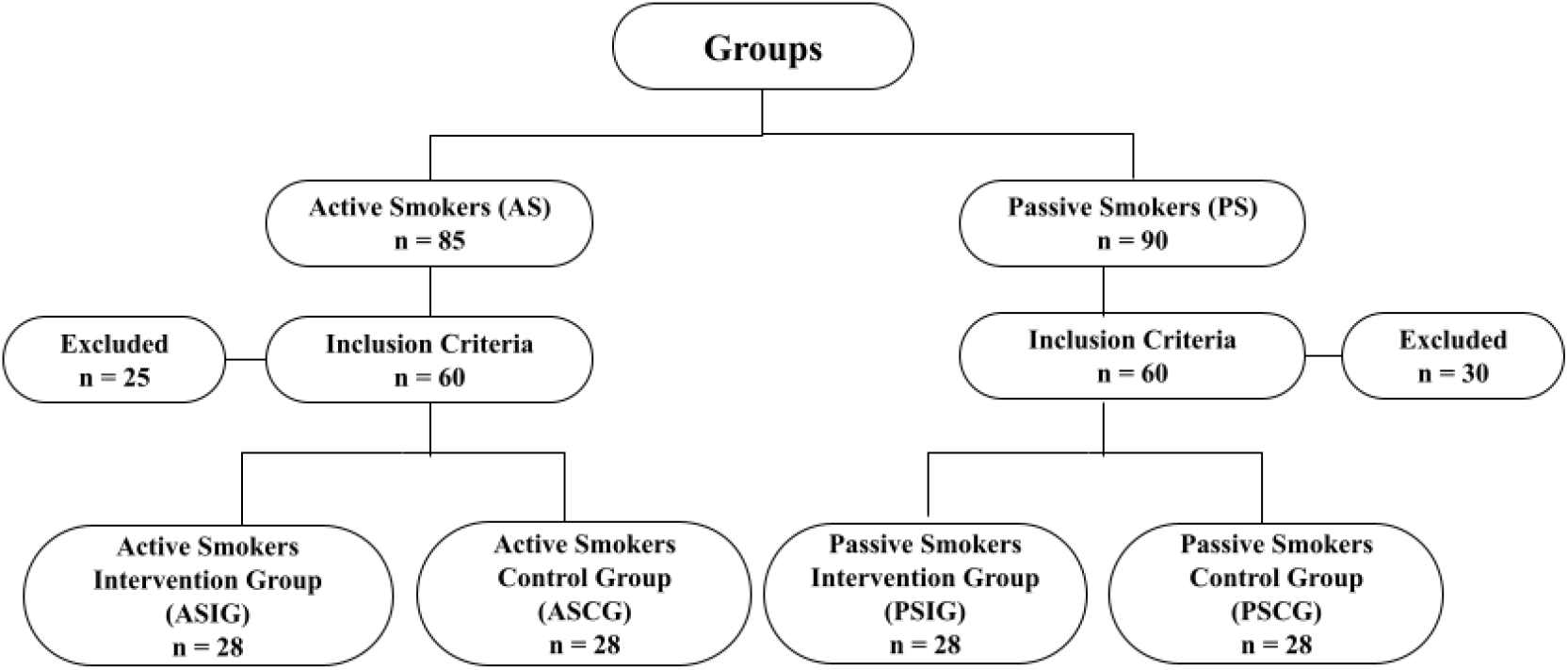
Study Flow Diagram.

#### Presentation and Seminars Program

During the presentations, the intervention groups were informed about the harmful effects of tobacco products as outlined in the literature. Particular emphasis was placed on the necessity of smoke-free zones for non-smokers, and the importance of smokers decreasing the number of cigarettes smoked. The seminars were held between 12:30 and 13:30 h.

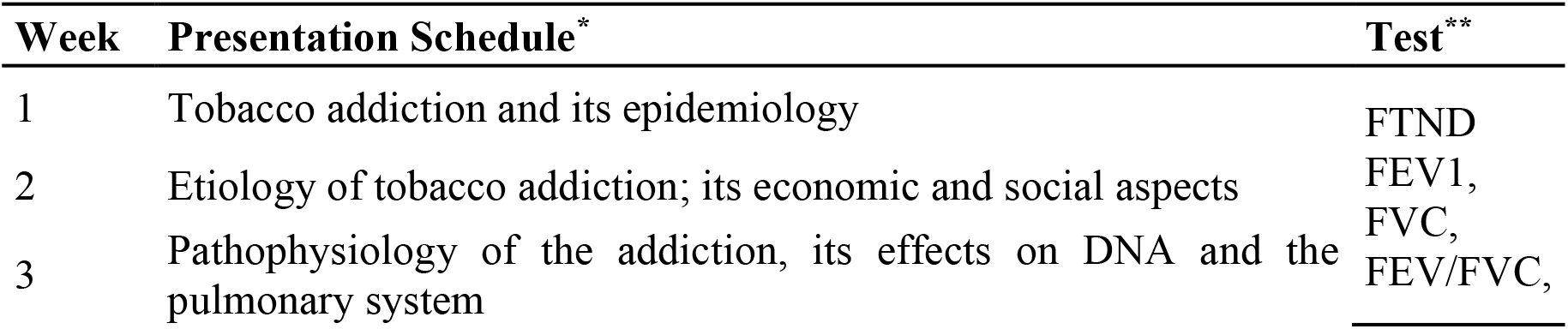
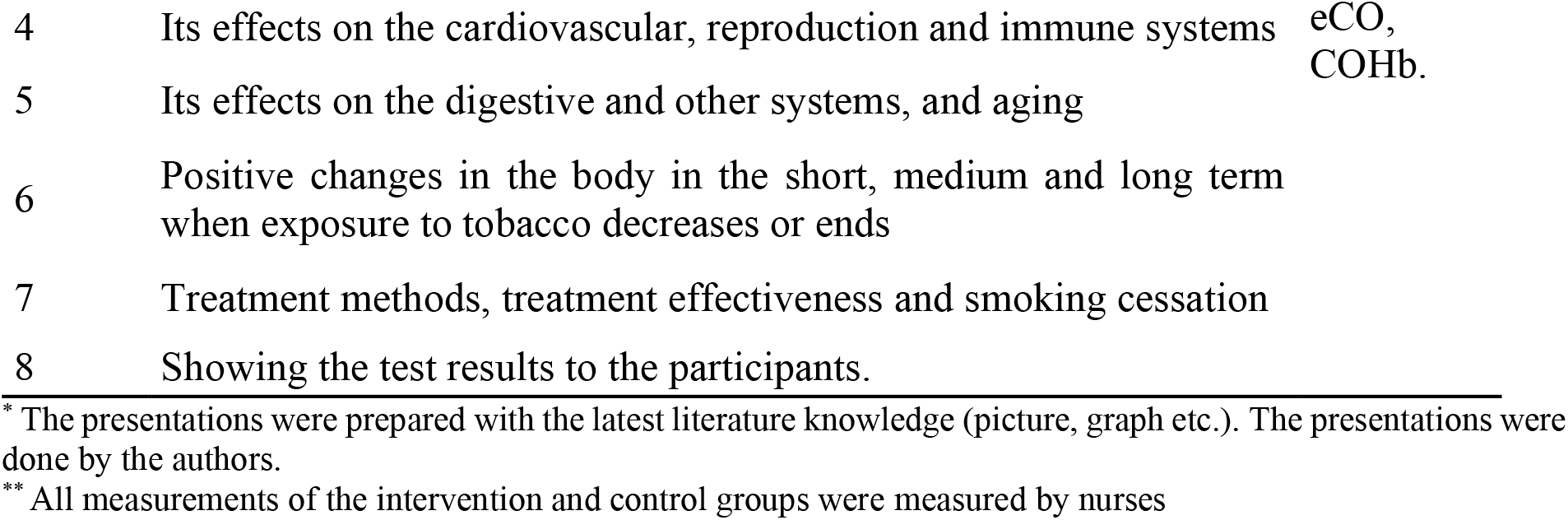

| Week | Presentation Schedule* | Test** |
| --- | --- | --- |
| 1 | Tobacco addiction and its epidemiology | FTND |
| 2 | Etiology of tobacco addiction; its economic and social aspects | FEV1,<br>FVC, |
| 3 | Pathophysiology of the addiction, its effects on DNA and the pulmonary system | FEV/FVC, |
| 4 | Its effects on the cardiovascular, reproduction and immune systems | eCO,<br>COHb. |
| 5 | Its effects on the digestive and other systems, and aging |  |
| 6 | Positive changes in the body in the short, medium and long term when exposure to tobacco decreases or ends |  |
| 7 | Treatment methods, treatment effectiveness and smoking cessation |  |
| 8 | Showing the test results to the participants. |  |

\* The presentations were prepared with the latest literature knowledge (picture, graph etc.). The presentations were done by the authors.
\*\* All measurements of the intervention and control groups were measured by nurses

### Data Collection and Tests

The data were collected from the participants using questionnaires and clinical tools. The questionnaires used were the sociodemographic questionnaire created by the authors. FTND. RFT, CO and COHb measurements were also conducted. All measurements of the intervention and control groups were made by nurses with ten years of experience in their field. In addition, the measurements of the subgroups were made in different places. The room in which the measurements were conducted was adjusted to optimum room temperature (22°C) and humidity (50-55%). Airflow and technical changes that can affect RFT were carefully monitored. The measurements were made at 16:00.

### FTND

This is a scale used to measure the severity of addiction (Uysal et al 2004). The highest score obtainable from the test is 10. Depending on the test score, ≤5 points, 5-6 points, and >7 points are considered as low, moderate and high levels of addiction, respectively.

### RFT (FEV_1_, FVC and FEV_1_/FVC)

RFT were measured with ZAN 100 Better Flow (Nspire Health GmbH, Germany). In the measurements, the criteria of the American Thoracic Society (ATS) were used (Miller et al 2005). FEV_1_ and FVC values were measured in liters/sec. The FEV_1_/FVC ratio was measured in percentage (%). The measurement device was calibrated by the authorized service before being applied.

### eCO and COHb

The parameters of eCO and COHb were measured using a Tabataba CO-Tester (FIM Médical, France). The eCO value was measured in parts per million (ppm). The COHb value was measured in percent (%). The measurement device was calibrated by the authorized service before being applied.

### Statistical Calculations and Ethical Committee Approval

The data was statistically analyzed using the SPSS.22 program (Chicago, IL). The results were shown as mean ± standard deviation 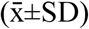, percentage (%) and number (n). The data were compared using the independent samples t-test and paired samples t-test. Differences were considered statistically significant for p<0.05 values. Ethical committee approval was obtained from the Ethics Committee of Selçuk University (2017/11).

## RESULTS

### Main Group Results (AS and PS Groups)

AS group (n = 60): Out of which were 21 females (35%) and were 39 males (65%). Out of which were 22 (36.7%) living in a dormitory and were 38 (63.3%) living in a house. PS group (n=60): Out of which were 38 females (%63.3) and 22 males (%36.7). Out of which were 16 (%26.7) living in a dormitory and were 44 (%73.3) living in a house. RFT, CO, COHb, and sociodemographic findings of the main groups are given in Tables 1 and 2.

**Table 1.** Sociodemographic findings of the main groups 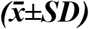.

| Group | Age | BMI | Income | Family Income | Households | Smokers* |
| --- | --- | --- | --- | --- | --- | --- |
| AS | 21.3±1.3 | 22.2±3.1 | 903.7±440.3 | 3846.7±1868.7 | 3±1.2 | 1.5±0.5 |
| PS | 20.3±1.4 | 22.6±3.9 | 787.5±316.2 | 2827.5±1083.6 | 2.9±0.8 | 1.4±0.5 |
| p | 0.001 | 0.491 | 0.1 | 0.02 | 0.416 | 0.732 |

Age: Years. BMI: kg/m<sup>2</sup>. Incomes: TL. Household: Person. \*Smokers: Number of smokers at house.

**Table 2:**
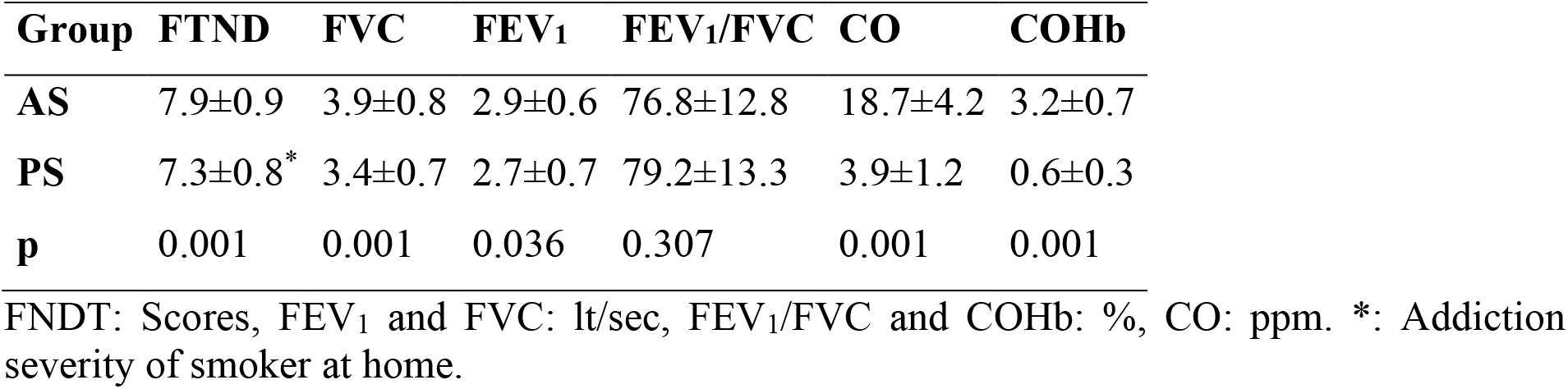
Test findings of the main groups 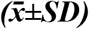.

### Sub-groups Results (Intervention and Control Groups)

ASIG consisted of 8 females (%26.7) and 22 males (%73.3). ASCG consisted of 13 females (%43.3) and 17 males (%56.7). PSIG consisted of 22 females (%73.3) and 8 males (%26.7). PSCG consisted of 16 females (%53.3) and 14 males (%46.7). 23 participants (%76.6) from ASIG living in a house and 7 participants (%23.4) living in a dormitory. 15 participants (%50) from ASCG living in a house and 15 participants (%50) living in a dormitory. 22 participants (%73.4) from PSIG and PSCG living in a house and 8 participants (%26.6) living in a dormitory.

The number of people in their house was: 3.2 ± 0.2 people in ASIG, 2.8 ± 0.2 people in ASCG, 2.8 ± 0.1 people in PSIG, and 2.9 ± 0.2 people in PSCG. In ASIG and ASCG, the number of smokers excluding them was calculated as 1.4 ± 0.9 and 1.5 ± 0.1 people (respectively). The number of people who exposed PSIG and PSCG tobacco smoke was calculated as 1.4 ± 0.1 in both groups.

When comparing the first and last weeks in ASIG, the difference between FTND, FEV_1_, CO, and COHb parameters was observed to be statistically significant (p<0.05). There was no significant difference between the first and last weeks of ASCG (p>0.05) (Table 3). The highest decrease in ASIG was in the FTND (↓%49.4) and CO (↓60%) parameter and the highest increase was in the FEV_1_(↑%10) parameter (Table 3).

**Table 3.** ASIG-ASCG comparison 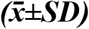.

| Variable | ASIG (n=30)* |  | ASCG (n=30)* |  | Group Comparison** |  |
| --- | --- | --- | --- | --- | --- | --- |
|  | 1<sup>th</sup> week | 8<sup>th</sup> week | 1<sup>th</sup> week | 8<sup>th</sup> week | 1 vs 1 | 8 vs 8 |
| FTND | 8.1±0.9 | 4.1±1.2 | 7.7±0.8 | 7.4±1.1 | | |
| p | **0.001** |  | 0.23 |  | 0.053 | **0.001** |
| FEV<sub>1</sub> | 3±0.5 | 3.3±0.5 | 2.8±0.7 | 2.9±0.6 | | |
| p | **0.027** |  | 0.76 |  | 0.225 | **0.006** |
| FVC | 4.1±0.9 | 4.2±0.6 | 3.7±0.7 | 3.6±0.7 | | |
| p | 0.45 |  | 0.81 |  | 0.058 | **0.001** |
| FEV<sub>1</sub>/FVC | 75.8±12.2 | 78.9±8.7 | 77.4±13.7 | 80.2±13.6 | | |
| p | 0.71 |  | 0.44 |  | 0.691 | 0.735 |
| CO | 18±4.2 | 7.2±2.1 | 19.4±4.2 | 20.3±4.2 | | |
| p | **0.001** |  | 0.39 |  | 0.212 | **0.001** |
| COHb | 3±0.7 | 1.8±0.5 | 3.5±0.7 | 3.6±0.6 | | |
| p | **0.001** |  | 0.66 |  | **0.006** | **0.001** |

FNNT: scores, FEV<sub>1</sub> and FVC: lt/sec, FEV<sub>1</sub>/FVC and COHb: %, CO: ppm. \* Paired Sample T Test. \*\* Student T Test.

There was a significant difference between FEV_1_, FVC, FEV_1_/FVC, and CO parameters of PSIG. The highest decrease in PSIG was in the CO parameter (↓%65.8) and the highest increase was in the FVC (↑%10) parameter (Table 4). There was no significant difference between the first and last weeks of ASCG and PSCG (Table 3 and 4, respectively). Other weeks’ measurements of ASIG and PSIG (between 2^nd^ and 7^th^ weeks) are given in Table 5.

**Table 4.**
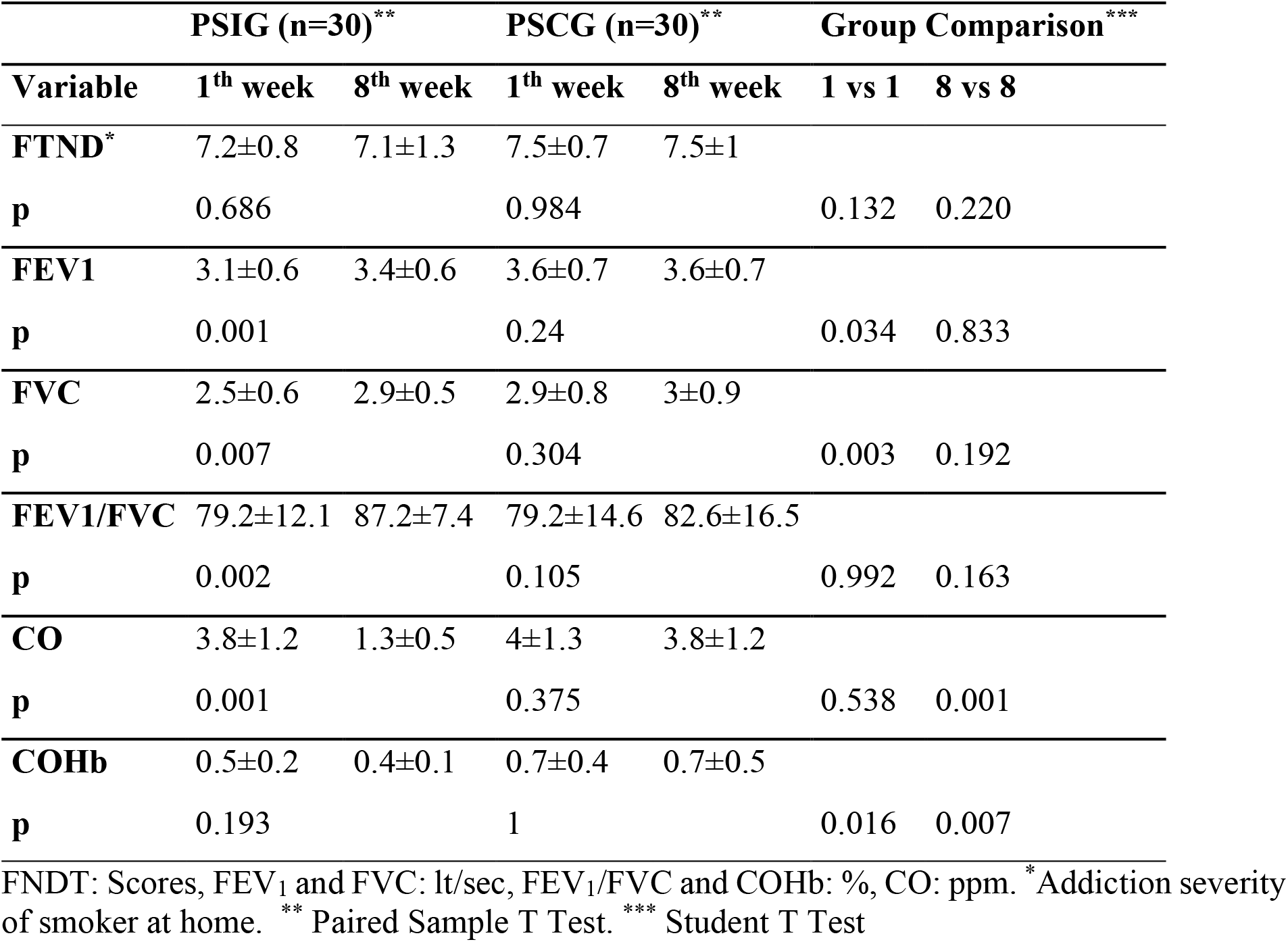
PSIG-PSCG comparison 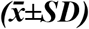.

**Table 5.** Findings between the 2nd-7th weeks of ASIG-PSIG 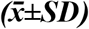.

| Group | Variable | 2<sup>nd</sup> week | 3<sup>th</sup> week | 4<sup>th</sup> week | 5<sup>th</sup> week | 6<sup>th</sup> week | 7<sup>th</sup> week |
| --- | --- | --- | --- | --- | --- | --- | --- |
| **ASIG** | **FTND** | 7.4±1 | 6.5±0.8 | 6.1±1.3 | 6±0.9 | 5.5±1 | 5.1±1 |
|  | **FVC** | 4±0.7 | 4±0.7 | 4.1±0.6 | 4.1±0.6 | 4.1±0.6 | 4.2±0.6 |
|  | **FEV** | 3±0.6 | 3.1±0.6 | 3.2±0.5 | 3.2±0.5 | 3.2±0.5 | 3.3±0.5 |
|  | **FEV<sub>1</sub>/FVC** | 76±14.2 | 78.2±12.2 | 79±9.9 | 78.7±8.9 | 78.4±8.5 | 79±8.8 |
|  | **CO** | 15.8±3.5 | 13.9±2.5 | 13.3±4.5 | 11.3±2.7 | 10.3±3.4 | 8.5±2.8 |
|  | **COHb** | 2.6±0.6 | 2.4±0.4 | 2.4±0.7 | 2.2±0.6 | 2.1±0.5 | 2±0,5 |
| **PSIG** | **FTND\*** | 7.4±0.7 | 6.9±0.9 | 7.1±0.9 | 7.2±1 | 7.3±1.21 | 7.27±1 |
|  | **FVC** | 3.3±0.8 | 3.2±0.7 | 3.3±0.6 | 3.3±0.6 | 3.4±0.6 | 3.4±0.6 |
|  | **FEV** | 2.7±0.6 | 2.7±0.7 | 2.9±0.5 | 2.9±0.5 | 2.9±0.5 | 2.9±0.5 |
|  | **FEV<sub>1</sub>/FVC** | 85.4±10.6 | 83.1±11.9 | 87.5±8.2 | 86.8±9.2 | 87.4±7.9 | 87.7±7.6 |
|  | **CO** | 3.7±0.9 | 2.5±0.9 | 2.4±0.8 | 2.1±0.9 | 1.9±0.8 | 1.4±0.6 |
|  | **COHb** | 0.6±0.1 | 0.4±0.2 | 0.4±0.2 | 0.5±0.1 | 0.5±0.1 | 0.5±0.1 |

FNNT: Scores. FEV<sub>1</sub> and FVC: lt/sec, FEV<sub>1</sub>/FVC and COHb: %, CO: ppm. Measurement of the 1<sup>th</sup> and 8<sup>th</sup> week of the intervention groups (ASIG and PSIG) were given in Table 3-4. \*: Addiction severity of smokers at home.

## DISCUSSION

If the problem of demand for tobacco and tobacco products is not resolved, a large number of people may die in the future due to tobacco-related diseases (Mathers and Loncar 2006). Various solutions and policies have been produced for the solution to this problem (Hyland et al. 2012, Vellios et al. 2018). The sustainable among these is to raise awareness of the harm of tobacco products. Various scientific researches have been conducted in the literature to raise awareness of tobacco products (Nyman et al. 2017, Park et al. 2018). However, in our literature review, it has been observed that intervention studies that both create long-term awareness and measure FTND, CO, COHb, and RFT are extremely limited. In our study, designed as an intervention research, active and passive smoker intervention groups were given a seminar/awareness about the harm of tobacco products for eight weeks. It is aimed to compare the findings obtained from the intervention groups with the control groups.

In our study, a statistical difference was detected parameters in ASIG (FTND, FEV_1_, CO, COHb) and PSIG (FEV_1_, FVC, FEV_1_/FVC, CO). No statistical difference was observed between FTND, RFT, CO, and COHb parameters in the control group (ASCG and PSCG) of both intervention groups. Previous studies have shown that awareness-raising by giving information and visuals about tobacco products may affect product use (not starting tobacco, quitting, reducing exposure, turning to the use of different tobacco products, etc.) (Mughal et al 2018, Yaddanapalli et al. 2018, Kim et al. 2018). The fact that the presented information and visuals cause positive changes in intervention groups is compatible with the literature. The fact that the presented information and visuals cause positive changes in ASIG and PSIG is compatible with the literature.

As a result of exposure to tobacco smoke, pathological conditions such as oxidative stress, inflammation, etc. may develop (Goel et al. 2017, Crotty Alexander et al. 2015, Liu et al. 2011). CO in the gas phase may play a role in the etiology of some diseases (George et al 2019). Exposure to high doses of CO may cause situations such as hypoxia, apoptosis, etc. (Hampson et al 2012). As long as exposure to both active and passive smoke continues, it can damage cells, tissues, and organs through hypoxia, apoptosis, and inflammation (Bhat et al. 2018). Especially these damages in the respiratory tract may cause decreases in RFT parameters (Allinson et al. 2017). Decreases in RFT parameters are used in the diagnosis of some diseases such as COPD (Vestbo et al 2011). These pathologies occurring in the body against tobacco smoke can be explained as a dose-response relationship (Schwartz and Cote 2016, Lugo et al. 2017). Thus, when both active and passive smokers are exposed to high doses, it is possible that tobacco-related diseases increase in proportion to the dose. For this reason, there are various studies about reducing the exposure dose and reducing the related diseases and smoking (Lee 2018; Saha et al. 2018; Leone et al. 2017). We believe that changes in intervention groups are related to the decrease of the FTND score (dose-response).

In addition, it may be possible to reduce various diseases (Asthma, COPD, etc.) that may occur in the future with a decrease in the FTND score. Another finding of the study was that the FTND score of the active smoker, in the environment where PSIG stay, was close to each other for 8 weeks. However, positive changes in some parameters of the group were found statistically significant. Tobacco smoke is an important air pollutant and can pollute the inhaled air (Rosen et al. 2015). When the passive smoker breathes side-stream smoke, it is possible that the situations that apply to the active smoker also apply to most passive smokers (Cao et al. 2015). For this reason, passive smokers may be affected by smoke even though they do not smoke, and tobacco-related diseases may occur (Zeng et al. 2012). Knowing the harm of tobacco smoke and staying away may be beneficial for the health of passive smokers (Ngo et al. 2020). We are of the opinion that PSIG has increased awareness with the current literature information and visuals, and they may protect themself despite being an active smoker in the same environment.

## CONCLUSION

Awareness programs may reduce the severity of addiction in active smokers and may help protect passive smokers. In the absence of any awareness, it may not positively change. As awareness increases, positive changes in some vital parameters may be possible. Up-to-date programs and policies are needed to make easy and sustainable awareness of both active and passive smoking.

### Limitations and Directions

The limitation of this study is that the study was conducted on young non-obese participants, the evaluation of the respiratory function test on the main parameters and the addiction severity of active smokers who exposed passive smokers to tobacco smoke was measured by passive smokers.

### Suggestions for Future Research

Research on the harms of tobacco should be focused on young people and childhood. The harm caused by second-hand and third-hand tobacco smoke to passive smokers should be emphasized. Tobacco addicts should participate in a program about possible situations before smoking cessation treatment.

### Main Points

1. High levels of carbon monoxide, carboxyhemoglobin and addiction severity can be reduced through awareness programs.
2. Through awareness programs, the burden of tobacco-related diseases can be reduced.
3. Long-term awareness of the harms of tobacco use can be beneficial.
4. Decreased vital parameters due to tobacco can be increased through awareness programs.

## Data Availability

.

